# STRENGTHENING RURAL HEALTH WORKFORCE DEVELOPMENT THROUGH ORGANIZATIONAL SUPPORT AND GENERATIONAL ADAPTATION: EVIDENCE FROM THE PHILIPPINES

**DOI:** 10.64898/2026.01.27.26344848

**Authors:** Fernan Torreno, Frincess Flores

## Abstract

**Introduction:** Rural and geographically isolated communities in the Philippines continue to face chronic shortages of healthcare professionals, particularly among younger generations who often prefer urban or overseas employment. Despite long-standing national deployment programs, many rural health units (RHUs) and barangay health stations (BHSs) still struggle to attract and retain competent staff. Understanding the organizational characteristics that support young professionals is essential for strengthening rural workforce sustainability in these settings. This study explored how rural health organizations in the Philippines adapt to generational expectations and support the next generation of healthcare workers in geographically isolated and disadvantaged areas (GIDAs).

**Methods:** A qualitative descriptive design was employed. Fourteen healthcare workers—physicians, nurses, midwives, nutritionists, barangay health workers, and administrative staff—were purposively selected from three GIDA municipalities in the Philippines. Semi-structured interviews were conducted in RHUs and BHSs, audio-recorded with consent, transcribed verbatim, anonymized, and analyzed using qualitative content analysis. Meaning units were coded and grouped into categories and themes through iterative team discussions to enhance credibility and confirmability.

**Results:** Five organizational characteristics emerged as essential for supporting young rural healthcare workers. Community-integrated mentorship enabled hands-on learning in barangay settings and strengthened cultural competence. Flexible and supportive work environments fostered teamwork, emotional safety, and shared responsibility. Clear pathways for professional growth—including access to training and opportunities for expanded roles and leadership—enhanced motivation and long-term commitment. Culturally grounded patient engagement improved community trust and the perceived relevance of health interventions. Finally, organizational adaptability to generational values, such as structured feedback, digital communication, and attention to work-life balance, was critical for retaining younger staff.

**Conclusion:** Rural health organizations that provide mentorship, supportive work environments, meaningful opportunities for professional growth, culturally grounded care, and generationally responsive practices are better positioned to attract and retain young healthcare professionals in GIDAs. These findings offer practical guidance for local government units, the Department of Health, and academic institutions in designing policies and programs that strengthen rural health workforce development in the Philippines.

## INTRODUCTION

Rural and geographically isolated communities in the Philippines have long faced persistent challenges in securing a stable, competent, and motivated health workforce. Despite decades of reforms, deployment programs, and investments in primary care, many rural health units (RHUs) and barangay health stations (BHS) continue to operate with chronic understaffing, limited professional support, and high turnover among young healthcare workers^1^. These shortages are particularly acute in geographically isolated and disadvantaged areas (GIDAs), where difficult terrain, limited transportation, and socioeconomic constraints further complicate the delivery of essential health services^2^. The consequences of these workforce gaps are profound: delayed care, fragmented continuity of services, increased burden on existing staff, and reduced community trust in the health system^3^.

The Philippine health sector has long recognized the importance of strengthening rural human resources for health (HRH). Programs such as the Doctors to the Barrios (DTTB), the Nurse Deployment Program (NDP), the Rural Health Midwives Program (RHMPP), and the broader Human Resources for Health (HRH) Deployment Program were designed to address shortages by assigning health professionals to underserved areas^4^. While these initiatives have improved access in many communities, they have not fully resolved the underlying issue of long term retention. Many young professionals complete their deployment contracts and subsequently transfer to urban hospitals, private clinics, or overseas employment^5^. This pattern reflects a broader generational shift in career aspirations, work life expectations, and professional identity among younger Filipino healthcare workers^6^.

Globally, rural health workforce shortages are not unique to the Philippines. Countries such as Papua New Guinea, Japan, Australia, and Canada face similar challenges in attracting and retaining young professionals in remote areas^7^. International literature consistently highlights the importance of mentorship, supportive supervision, community immersion, and positive rural exposure in shaping rural career intentions^8^. However, the Philippine context presents unique structural and cultural dynamics. The decentralization of health services under the Local Government Code of 1991 places significant responsibility on local government units (LGUs) to manage health personnel, often resulting in variability in resources, leadership styles, and organizational cultures across municipalities^9^. These differences influence the experiences of young healthcare workers and shape their decisions to stay or leave.

Another emerging factor is the generational transition within the health workforce. Many rural health organizations were designed around the expectations and working styles of older generations—Baby Boomers and Generation X—who often valued hierarchy, long service, and traditional supervisory structures^10^. In contrast, Generation Z, now entering the workforce, prioritizes mentorship, digital communication, structured feedback, mental well being, and opportunities for rapid professional growth^11^. They seek workplaces that are collaborative, technologically enabled, and aligned with their personal values^12^. Understanding how rural health organizations can adapt to these generational preferences is essential for building a sustainable workforce.

Despite the importance of these issues, there remains limited empirical research examining the organizational characteristics that support the recruitment, motivation, and retention of young healthcare professionals in Philippine rural settings. Existing studies often focus on individual motivations, deployment program evaluations, or broad health system challenges^13^. Few studies explore the internal dynamics of rural health organizations—their culture, leadership, mentorship structures, and adaptability to generational change^14^. This gap limits the ability of policymakers, LGUs, and academic institutions to design targeted interventions that address the needs of the next generation of rural health workers.

The Philippine rural health system operates within a complex interplay of local governance, community expectations, and resource constraints. RHUs and BHS facilities often function with limited budgets, outdated infrastructure, and insufficient staffing patterns^15^. Young healthcare workers entering these environments frequently encounter challenges such as heavy workloads, limited supervision, and unclear career pathways^16^. These conditions can contribute to early burnout, reduced job satisfaction, and decisions to seek employment elsewhere^17^. At the same time, many rural communities rely heavily on their local health workers, viewing them not only as service providers but also as trusted community members^18^. This dual role can be both rewarding and demanding for young professionals who are still developing their clinical confidence and professional identity.

Mentorship has been identified as a critical factor in supporting young healthcare workers in rural settings. Studies from various countries show that structured mentorship programs enhance clinical competence, build confidence, and foster a sense of belonging within the organization^19^. In the Philippine context, mentorship often occurs informally through senior staff, barangay health workers, or municipal health officers who guide younger colleagues during community visits, immunization campaigns, and maternal and child health activities^20^. However, the availability and quality of mentorship vary widely across LGUs, depending on leadership commitment, staffing levels, and organizational culture^21^.

Organizational culture plays a significant role in shaping the experiences of young healthcare workers. Supportive environments characterized by teamwork, open communication, and shared responsibility can mitigate the challenges of working in resource limited settings^22^. Conversely, rigid hierarchies, poor communication, and lack of recognition can exacerbate stress and contribute to turnover^23^. Generation Z workers, in particular, value psychological safety, collaborative decision making, and opportunities to contribute meaningfully to organizational goals^24^. Rural health organizations that fail to adapt to these expectations may struggle to retain young staff.

Professional growth opportunities are another key factor influencing retention. Young healthcare workers often seek continuous learning, specialization, and leadership roles^25^. In rural areas, however, access to training programs, scholarships, and career advancement pathways may be limited^26. Some LGUs support their staff through training budgets or partnerships with academic institutions, while others lack the resources to do so^27^. This uneven distribution of opportunities contributes to disparities in workforce stability across regions.

Cultural competence is also essential in rural health practice. Many rural communities in the Philippines have distinct cultural norms, languages, and traditions that influence health seeking behaviors^28^. Young healthcare workers who are unfamiliar with these cultural contexts may initially struggle to build rapport with patients^29^. Organizational support—such as orientation programs, community immersion activities, and guidance from experienced staff—can help young professionals develop the cultural sensitivity needed for effective practice^30^.

Finally, the rapid digitalization of healthcare presents both opportunities and challenges for rural health organizations. Generation Z workers are highly comfortable with technology and expect digital tools to support communication, documentation, and service delivery_31_. However, many rural facilities lack reliable internet connectivity, updated equipment, or electronic health systems^32^. Organizations that invest in digital infrastructure may be better positioned to attract and retain young professionals who value efficiency and technological integration^33^.

Given these complexities, there is a pressing need to understand how rural health organizations in the Philippines can adapt to the expectations of the next generation of healthcare workers. This study addresses this need by exploring the organizational characteristics that support young professionals in rural communities in Northern Luzon. Using a qualitative descriptive design, the study captures the lived experiences of multi professional healthcare workers—including physicians, nurses, midwives, nutritionists, barangay health workers, and administrative staff—who work in GIDAs. Their insights provide a grounded understanding of what helps young professionals thrive, what challenges they face, and what organizational practices contribute to long term retention.

By examining these organizational characteristics, the study contributes to a deeper understanding of rural workforce sustainability in the Philippines. The findings offer practical implications for LGUs, the Department of Health, academic institutions, and policymakers seeking to strengthen rural health systems. Ultimately, supporting the next generation of rural healthcare workers is essential for achieving equitable access to quality healthcare across the country and advancing the goals of Universal Health Care.

## METHODS

### Study Design

This study employed a qualitative descriptive design, an approach widely used in health services research to capture participants’ experiences in a manner that remains close to their natural language and everyday understanding^1. Qualitative description is particularly appropriate when the goal is to generate practical insights for policy and organizational improvement rather than to develop or test theory^2. It allows researchers to explore phenomena in depth while maintaining a low level of inference, making it suitable for examining the organizational characteristics that support young healthcare workers in rural settings^3^. The design aligns with the study’s aim of understanding how rural health organizations in the Philippines adapt to generational expectations and support the next generation of healthcare professionals.

### Study Setting

The study was conducted in three geographically isolated and disadvantaged areas (GIDAs) in the Philippines. These municipalities were selected because they represent typical rural contexts characterized by limited transportation, mountainous terrain, and constrained access to secondary and tertiary health facilities^4^. Each municipality operates a Rural Health Unit (RHU) as the primary provider of public health services, supported by several Barangay Health Stations (BHS) staffed by midwives and barangay health workers (BHWs)^5^.

The selected sites vary in population size, socioeconomic conditions, and local government capacity, reflecting the diversity of rural health environments in the region. All three municipalities rely heavily on national deployment programs and contractual staff to maintain essential services^6^. Internet connectivity is intermittent, and health information systems are largely paper based, although some facilities have begun adopting digital tools for reporting and communication^7^. These contextual factors shape the organizational dynamics experienced by young healthcare workers and provide a rich environment for exploring the study’s research questions.

### Participants and Sampling

A purposive sampling strategy was used to recruit participants who could provide rich, relevant, and diverse perspectives on the organizational characteristics of rural health facilities^8^. Fourteen healthcare workers participated in the study, representing a range of professional roles: physicians, nurses, midwives, nutritionists, barangay health workers, and administrative staff. This multi professional composition allowed the study to capture the experiences of both formally trained health professionals and community based workers who play critical roles in rural health delivery^9^.

Inclusion criteria were:

1. Currently employed in an RHU or BHS within the selected GIDAs.
2. At least six months of work experience in the rural setting.
3. Willingness to participate in an in depth interview.
4. Ability to communicate in English, Filipino, or Ilocano.

Exclusion criteria included temporary volunteers or students on short term immersion programs, as their experiences may not reflect sustained organizational engagement^10^.

The sample size was guided by the principle of information power, which suggests that smaller samples are sufficient when the study aim is narrow, the sample is specific, and the quality of dialogue is strong^11^. Recruitment continued until the research team determined that no new insights were emerging, indicating data sufficiency rather than strict saturation^12^.

### Data Collection

Data were collected through semi structured, face to face interviews conducted between January and March. Interviews took place in private rooms within RHUs or BHS facilities to ensure confidentiality and minimize disruptions. Each interview lasted between 45 and 75 minutes.

An interview guide was developed based on existing literature on rural health workforce retention, organizational culture, and generational expectations^13^. The guide included open ended questions exploring:

- Experiences working in rural health facilities
- Perceptions of organizational support
- Mentorship and supervision
- Opportunities for professional growth
- Workplace culture and teamwork
- Adaptation to generational needs
- Challenges and motivations for staying or leaving

Probing questions were used to encourage elaboration and clarify meanings^14^. The guide was pilot tested with two rural health workers outside the study sites to ensure clarity and relevance; minor adjustments were made accordingly^15^.

All interviews were audio recorded with participants’ consent. Field notes were taken to capture non verbal cues, contextual details, and initial analytic impressions^16^. Interviews conducted in Filipino or Ilocano were translated into English by a bilingual researcher and verified by another team member to ensure accuracy^17^.

## Researcher Positionality

The primary researcher has professional experience in rural health systems and academic training in qualitative research. This background facilitated rapport with participants but also required reflexivity to minimize potential bias^18^. Throughout the study, the researcher maintained a reflexive journal documenting assumptions, decisions, and emotional responses during data collection and analysis^19^. Regular discussions with the research team helped ensure that interpretations remained grounded in participants’ accounts rather than researcher expectations^20^.

### Data Management and Transcription

Audio recordings were transcribed verbatim by trained research assistants. Transcripts were reviewed while listening to the recordings to ensure accuracy^21^. All identifying information—names, locations, and specific events—was removed or replaced with pseudonyms to protect confidentiality^22^. Digital files were stored on password protected devices accessible only to the research team^23^.

### Data Analysis

Data were analyzed using qualitative content analysis, a systematic method for identifying patterns and categories within textual data^24^. The analysis followed the steps outlined by Graneheim and Lundman^25^:

1. Familiarization: Researchers read each transcript multiple times to gain an overall understanding of the content.
2. Identification of Meaning Units: Segments of text related to the research questions were highlighted as meaning units^26^.
3. Condensation and Coding: Meaning units were condensed while preserving core meanings, then assigned descriptive codes^27^.
4. Categorization: Codes were grouped into categories based on similarities and differences^28^.
5. Abstraction: Categories were further refined into overarching themes representing the organizational characteristics that support young rural healthcare workers^29^.

NVivo software was used to organize data, track coding decisions, and facilitate retrieval of coded segments^30^. Coding was conducted independently by two researchers, and discrepancies were resolved through discussion to enhance analytic rigor^31^.

### Ensuring Trustworthiness

The study adhered to established criteria for trustworthiness in qualitative research: credibility, dependability, confirmability, and transferability^32^.

### Credibility

Credibility was enhanced through prolonged engagement in the field, triangulation of perspectives across professional roles, and member checking^33^. Participants were invited to review summaries of their interviews to verify accuracy and provide clarifications^34^.

### Dependability

An audit trail documenting methodological decisions, coding processes, and analytic memos was maintained throughout the study^35^. Regular team meetings ensured consistency in coding and interpretation^36^.

### Confirmability

Reflexive journaling and peer debriefing helped minimize researcher bias^37^. All interpretations were grounded in participants’ direct statements, supported by verbatim quotations^38^.

### Transferability

Thick descriptions of the study setting, participant characteristics, and organizational context allow readers to assess the applicability of findings to other rural settings^39^.

### Ethical Considerations

Ethical approval was obtained from the Philippine Region 1 Ethical Review Committee (Protocol No. NURDD826A 2026). All participants provided written informed consent prior to data collection. They were informed of their right to withdraw at any time without consequences^40^. Confidentiality was ensured through anonymization of transcripts and secure data storage^41^.

### Rigor and Reflexivity

The research team engaged in continuous reflexivity to ensure that interpretations remained grounded in participants’ experiences rather than researcher assumptions^42^. Reflexive discussions were held after each interview and during coding sessions to examine how personal backgrounds, professional experiences, and expectations might influence the analysis^43^.

### Summary of Methodological Strengths

This study’s methodological strengths include its multi professional sample, in depth interviews, rigorous analytic procedures, and strong attention to trustworthiness. The use of qualitative description allowed for a nuanced yet practical understanding of organizational characteristics relevant to rural health workforce development^44^.

## RESULTS

A total of fourteen healthcare workers participated in the study, representing physicians, nurses, midwives, nutritionists, barangay health workers (BHWs), and administrative staff. Participants ranged from 22 to 41 years old, with most belonging to the younger segment of the rural health workforce. Their years of service varied from 6 months to 12 years, reflecting a mix of early career and moderately experienced professionals. As shown in Table 1, the sample provided a diverse representation of roles across three geographically isolated and disadvantaged areas (GIDAs) in Northern Luzon.

**Table 1.** Demographic and professional profile of participants (N = 14)

| Participant code | Age group (years) | Sex | Profession | Years in rural service | Facility type |
| --- | --- | --- | --- | --- | --- |
| P01 | 20–24 | F | Nurse | 0–2 | Rural Health Unit (RHU) |
| P02 | 25–29 | F | Midwife | 6–9 | Barangay Health Station (BHS) |
| P03 | 20–24 | F | Barangay health worker (BHW) | 0–2 | BHS |
| P04 | 30–34 | M | Physician | 3–5 | RHU |
| P05 | 25–29 | F | Nutritionist | 3–5 | RHU |
| P06 | 40–44 | F | Midwife | 10+ | BHS |
| P07 | 25–29 | M | Nurse | 0–2 | RHU |
| P08 | 30–34 | F | Administrative staff | 3–5 | RHU |
| P09 | 25–29 | F | Nurse | 0–2 | BHS |
| P10 | 35–39 | M | Physician | 6–9 | RHU |
| P11 | 20–24 | F | BHW | 0–2 | BHS |
| P12 | 25–29 | F | Midwife | 6–9 | BHS |
| P13 | 30–34 | M | Nutritionist | 3–5 | RHU |
| P14 | 20–24 | F | Nurse | 0–2 | RHU |

Table 1 presents the demographic and professional profile of the 14 primary health care providers who participated in the study, highlighting a range of ages, professions, and facility types across the rural settings. The table functions as qualitative participant characteristics display and uses de-identified participant codes (P01–P14), which is consistent with recommended reporting practices.

Analysis of the interview data generated five major themes that describe the organizational characteristics supporting young healthcare workers in rural settings:

1. Community integrated mentorship;
2. Flexible and supportive work environments;
3. Clear pathways for professional growth;
4. Culturally grounded patient engagement;
5. Organizational adaptability to generational values.

To illustrate how these themes interrelate, Figure 1 presents a thematic map showing the central role of organizational support in shaping young workers’ motivation and retention.

**Figure 1.**
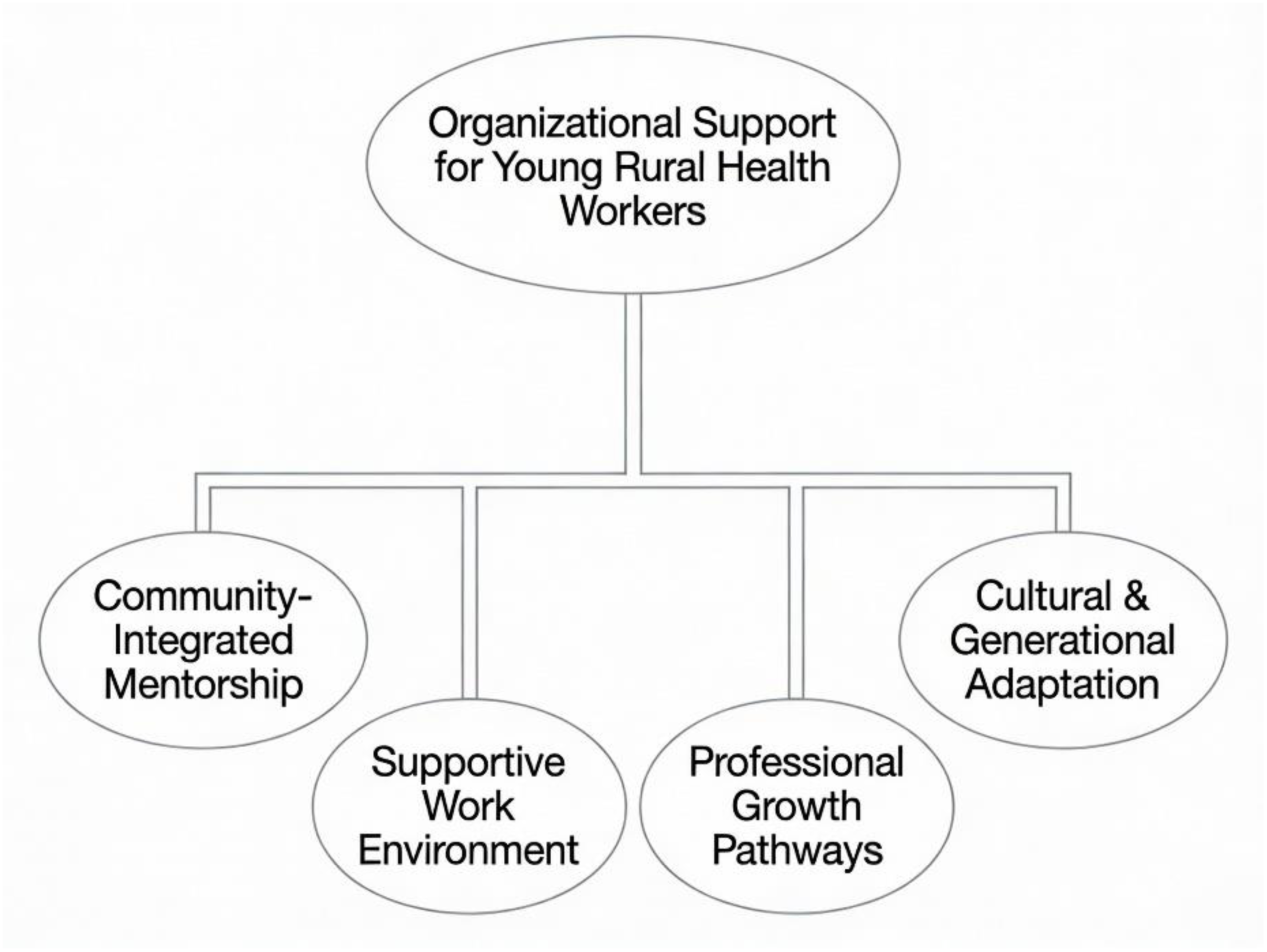
Thematic Map of Organizational Characteristics

Figure 1 depicts the overarching theme “Organizational Support for Young Rural Health Workers” at the center-top. Four main themes branch downward: Community-Integrated Mentorship, Supportive Work Environment, Professional Growth Pathways, and Cultural & Generational Adaptation. This thematic map illustrates key organizational factors supporting young rural health providers A summary of the themes and sub themes is presented in Table 2.

**Table 2.** Themes and Sub Themes.

| Theme | Sub-themes |
| --- | --- |
| 1. Community-Integrated Mentorship | Hands-on learning; Senior staff guidance; Community immersion |
| 2. Flexible and Supportive Work Environments | Teamwork; Emotional safety; Leadership approachability |
| 3. Clear Pathways for Professional Growth | Training access; Expanded roles; Leadership opportunities |
| 4. Culturally Grounded Patient Engagement | Respect for elders; Local language use; Community trust-building |
| 5. Organizational Adaptability to Generational Values | Digital communication; Work-life balance; Structured feedback |

### Themes and sub-themes of organizational characteristics

#### Theme 1: Community Integrated Mentorship

Participants consistently emphasized that mentorship in rural health settings is embedded in community activities rather than formal training structures. Young healthcare workers described barangay visits, home based consultations, and community campaigns as essential learning environments that strengthened their confidence and cultural competence^1.

A nurse explained:

> “During barangay visits, the midwife would guide me—how to talk to mothers, how to assess malnourished children. That’s where I really learned.”

Senior midwives and BHWs, many with decades of experience, served as accessible mentors who provided practical advice and emotional support. Their guidance helped young workers navigate local customs, political dynamics, and community expectations.

This theme highlights that mentorship is not a program but a lived practice, deeply embedded in the rhythms of rural health work.

#### Theme 2: Flexible and Supportive Work Environments

Participants described their workplaces as supportive when teamwork, open communication, and shared responsibility were present. Younger workers valued environments where they felt safe to ask questions, admit mistakes, and seek help^2^.

One participant shared:

> “Here, we help each other. If someone is overwhelmed, we step in. That’s why I feel comfortable staying.”

Supportive leadership—particularly municipal health officers who modeled approachability and fairness—was repeatedly cited as a factor that reduced stress and strengthened commitment.

However, some participants noted inconsistencies across facilities, with certain workplaces exhibiting rigid hierarchies that discouraged initiative.

#### Theme 3: Clear Pathways for Professional Growth

Opportunities for training, specialization, and leadership roles were central to young workers’ motivation. Participants expressed strong interest in continuous learning but noted that access to training varied depending on LGU resources and leadership priorities^3^.

Those who received support for seminars, scholarships, or expanded responsibilities described feeling more valued and more likely to stay.

One nurse explained:

> “When they send me to trainings, I feel they believe in me. It makes me want to stay longer.”

Conversely, limited opportunities contributed to frustration and thoughts of transferring to urban or overseas positions.

#### Theme 4: Culturally Grounded Patient Engagement

Participants emphasized that effective rural health practice requires deep cultural sensitivity, especially in communities where traditional beliefs, local dialects, and long standing social norms shape health seeking behaviors^4. Many young healthcare workers initially struggled with these nuances but gradually developed confidence through repeated exposure and mentorship.

One midwife explained:

> “You cannot just enter a barangay and expect people to follow you. You need to understand their customs, their language, their way of talking. Respect is everything.”

Participants described how elders in the community often expect a particular tone of politeness, slower pacing of explanations, and acknowledgment of local traditions. Younger workers learned to adjust their communication style accordingly.

Another participant shared:

> “At first, I was too direct. The older patients didn’t like it. My senior taught me how to soften my approach, how to listen more. That changed everything.”

Cultural competence was not viewed as an optional skill but as a core requirement for building trust. Participants noted that once trust was established, community members became more receptive to health advice, more consistent in follow ups, and more willing to collaborate during public health campaigns.

This theme highlights that organizational support for cultural learning—such as community immersion, mentorship, and orientation programs—plays a crucial role in helping young workers adapt and thrive.

#### Theme 5: Organizational Adaptability to Generational Values

Participants consistently described generational differences in expectations, communication styles, and work life priorities. Younger healthcare workers—particularly those from Generation Z—valued digital communication, structured feedback, and clear boundaries between work and personal life^5^.

One young nurse explained:

> “We like feedback. We want to know if we’re doing things right. Not after six months—right away.”

Another participant emphasized the importance of digital tools:

> “We use group chats for coordination. It’s faster. But some older staff prefer paper memos. That causes delays.”

Participants also noted that younger workers value work life balance, which sometimes clashes with traditional expectations of long hours and unquestioned availability. Organizations that recognized these preferences—by offering flexible schedules, acknowledging mental health needs, or modernizing communication systems—were perceived as more supportive and attractive to young professionals.

Senior staff also described the need for intergenerational coexistence, echoing findings from other rural health studies^6^. They recognized that adapting teaching styles, communication patterns, and expectations was essential for sustaining a harmonious and effective workforce.

This theme underscores that organizational adaptability is not merely beneficial but necessary for retaining the next generation of rural healthcare workers. To further illustrate these themes, Table 3 presents representative quotations from young rural health workers alongside their professional roles.

**Table 3.** Representative Quotations per Theme.

| Theme | Representative quotation | Profession |
| --- | --- | --- |
| 1. Community-Integrated Mentorship | “The midwives taught me things I never learned in school—how to talk to elders, how to negotiate with barangay captains.” | Nurse |
| 2. Flexible and Supportive Work Environments | “Here, we help each other. If someone is overwhelmed, we step in. That’s why I feel comfortable staying.” | Physician |
| 3. Clear Pathways for Professional Growth | “When they send me to trainings, I feel they believe in me. It makes me want to stay longer.” | Nutritionist |
| 4. Culturally Grounded Patient Engagement | “You need to understand their customs. Respect is everything in the barangay.” | Midwife |
| 5. Organizational Adaptability to Generational Values | “We like real-time feedback. It helps us improve faster.” | Nurse |

Building on these themes, Figure 2 presents a conceptual model illustrating how organizational structures shape supportive processes and, ultimately, the outcomes of young rural healthcare workers.

**Figure 2.**
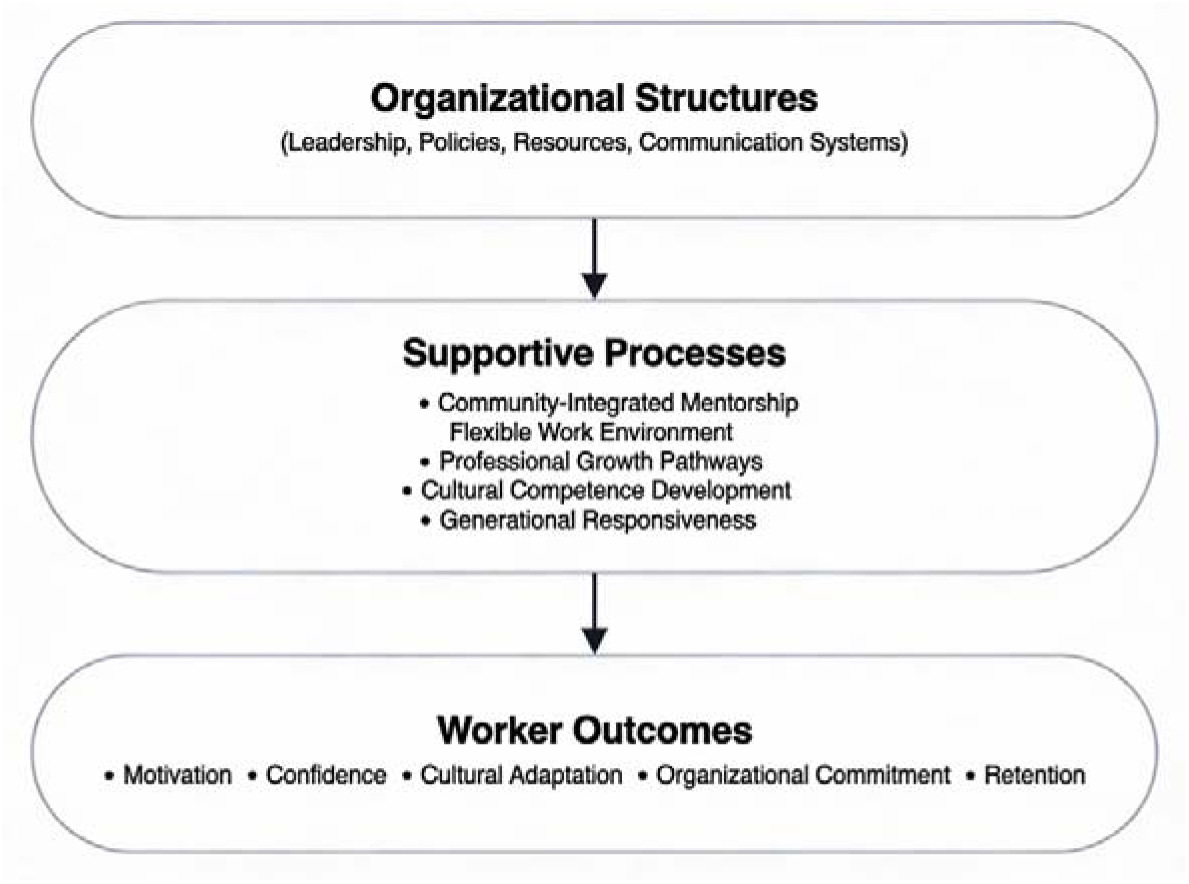
Conceptual Model of Organizational Support for Young Rural Healthcare Workers

### Integrative Summary

The findings of this study reveal that young healthcare workers in rural Philippine settings thrive when organizational structures and practices align with their developmental needs, cultural context, and generational expectations. Community integrated mentorship provides practical and emotional support, while flexible work environments foster psychological safety and teamwork. Clear pathways for professional growth enhance motivation and long term commitment. Culturally grounded engagement strengthens community trust, and organizational adaptability to generational values ensures that young workers feel understood and supported.

Together, these characteristics form a holistic organizational ecosystem that nurtures the next generation of rural healthcare professionals. The thematic map and conceptual model illustrate how these elements interact to shape worker experiences and influence retention. These insights offer practical implications for local government units, health administrators, and policymakers seeking to strengthen rural health systems and build a sustainable workforce.

## DISCUSSION

This study explored the organizational characteristics that support young healthcare workers in rural and geographically isolated and disadvantaged areas (GIDAs) in the Philippines. Through in depth interviews with fourteen early career professionals, five interrelated themes emerged: community integrated mentorship, flexible and supportive work environments, clear pathways for professional growth, culturally grounded patient engagement, and organizational adaptability to generational values. Together, these findings illuminate how rural health organizations can cultivate a sustainable and motivated workforce capable of addressing persistent shortages in underserved areas. The discussion below situates these findings within existing literature, highlights their contributions, and outlines implications for policy and practice.

### Community Integrated Mentorship as a Foundation for Rural Workforce Development

The first theme—community integrated mentorship—underscores the centrality of experiential, relationship based learning in rural health settings. Participants consistently described mentorship not as a formal program but as an organic, community embedded process shaped by barangay visits, home based consultations, and collaborative public health activities. This aligns with findings from Japan, where rural healthcare professionals emphasized the importance of teamwork, communication, and role modeling in nurturing the next generation^1. Similarly, global studies highlight that exposure to rural practice, supportive supervision, and positive role models are key determinants of young professionals’ willingness to remain in rural areas^2^.

The present study extends this literature by demonstrating that mentorship in the Philippine rural context is inseparable from community immersion. Senior midwives and barangay health workers—many with decades of experience—serve as cultural brokers who guide younger staff through local norms, political dynamics, and community expectations. This relational mentorship fosters not only clinical competence but also cultural fluency, confidence, and a sense of belonging. These findings reinforce the argument that rural health systems must invest in structured yet flexible mentorship models that leverage the expertise of long serving local staff while supporting the learning needs of younger workers.

### Supportive Work Environments and the Importance of Psychological Safety

The second theme highlights the role of supportive work environments in shaping young workers’ motivation and retention. Participants valued workplaces characterized by teamwork, open communication, and shared responsibility. These findings echo earlier research showing that respectful supervisors, excellent role models, and useful feedback are essential components of effective clinical supervision in rural settings^1^. International studies similarly emphasize that supportive organizational cultures—where staff feel safe to ask questions, admit mistakes, and seek help—are critical for sustaining rural health workforces^2^.

In the present study, psychological safety emerged as a key determinant of job satisfaction. Young workers described feeling more confident and committed when leaders were approachable and when colleagues collaborated during high workload periods. Conversely, rigid hierarchies and punitive communication styles discouraged initiative and contributed to thoughts of leaving. These insights suggest that leadership development programs for rural health managers should prioritize relational competencies, including empathy, communication, and conflict resolution.

Moreover, the variability in workplace culture across municipalities indicates that supportive environments are not guaranteed. Strengthening organizational climate requires intentional policy interventions, such as standardized orientation programs, team building activities, and leadership coaching. These strategies can help ensure that supportive work environments are not dependent solely on individual personalities but are embedded in organizational norms.

### Professional Growth Opportunities as Drivers of Motivation and Retention

The third theme—clear pathways for professional growth—highlights the importance of continuous learning and career development in retaining young rural healthcare workers. Participants expressed strong interest in training, specialization, and leadership roles, but access to these opportunities varied widely depending on local government unit (LGU) resources and leadership priorities. This mirrors findings from Australia, where young rural practitioners emphasized the need for structured learning environments and supportive guidance to sustain motivation^3^.

The present study contributes to this discourse by showing that professional growth is not merely a personal aspiration but a relational and organizational process. When young workers were supported to attend seminars, pursue certifications, or take on expanded responsibilities, they interpreted these opportunities as signs of trust and investment. This sense of being valued strengthened their commitment to the organization and reduced their desire to migrate to urban or overseas positions.

Conversely, limited access to training created frustration and a sense of stagnation. These findings underscore the need for national and local policies that ensure equitable access to professional development across rural areas. Potential strategies include scholarship programs, remote learning platforms, rotational training schemes, and partnerships with academic institutions. Investing in professional growth is not only beneficial for individual workers but also essential for improving the quality of rural health services.

### Cultural Competence as a Core Component of Rural Practice

The fourth theme emphasizes the importance of culturally grounded patient engagement in rural health work. Participants described how understanding local customs, dialects, and social norms was essential for building trust and delivering effective care. This aligns with findings from Japan, where professionalism and courtesy toward older adults were identified as critical characteristics for rural healthcare providers^1. Similarly, studies from Canada and Australia highlight that familiarity with rural community life and cultural expectations influences both patient relationships and career satisfaction^4^.

The present study deepens this understanding by illustrating how cultural competence is learned through mentorship and community immersion. Young workers initially struggled with communication styles and expectations of politeness, particularly when interacting with elders. Through guidance from senior staff, they learned to adjust their tone, pacing, and approach. This process of cultural adaptation strengthened their relationships with patients and enhanced their effectiveness in public health campaigns.

These findings suggest that cultural competence should be explicitly integrated into rural health training programs. Orientation modules, community immersion activities, and mentorship from local staff can help young workers navigate cultural complexities more effectively. Moreover, recognizing cultural competence as a core skill—not an optional attribute—can strengthen rural health systems’ responsiveness to community needs.

### Generational Adaptation as a Requirement for Sustainable Rural Health Systems

The fifth theme—organizational adaptability to generational values—highlights the evolving expectations of younger healthcare workers, particularly those from Generation Z. Participants emphasized their preference for digital communication, structured feedback, and work life balance. These findings resonate with research showing that younger generations value pragmatic thinking, real time feedback, and efficient work processes^5. Studies from Japan further indicate that young physicians increasingly prioritize work life balance and avoid specialties perceived as demanding or uncontrollable^6^.

In the present study, generational differences created both opportunities and tensions. Younger workers introduced digital tools, such as messaging apps, that improved coordination and efficiency. However, older staff sometimes resisted these changes, preferring traditional communication methods. Similarly, younger workers’ desire for clear boundaries and mental health considerations occasionally clashed with expectations of long hours and unquestioned availability.

These dynamics underscore the need for intergenerational dialogue and organizational flexibility. Rural health systems must adapt to the preferences of younger workers while preserving the wisdom and experience of older staff. Strategies may include hybrid communication systems, structured feedback mechanisms, flexible scheduling, and leadership training focused on intergenerational management. By embracing generational diversity, organizations can create more inclusive and sustainable work environments.

### Interconnectedness of Themes and Implications for Rural Health Systems

Although the five themes are presented separately, they are deeply interconnected. Community integrated mentorship supports cultural competence; supportive work environments enhance professional growth; and generational adaptability strengthens mentorship relationships. Together, these elements form a holistic organizational ecosystem that nurtures young rural healthcare workers.

The conceptual model developed in this study illustrates how organizational structures (leadership, policies, resources) shape supportive processes (mentorship, teamwork, cultural learning), which in turn influence worker outcomes (motivation, confidence, retention). This model aligns with global frameworks emphasizing the importance of integrated, context responsive strategies for rural workforce sustainability^2^.

For policymakers, these findings highlight the need for multi level interventions that address both structural and relational dimensions of rural health work. Investments in infrastructure and training must be complemented by efforts to strengthen organizational culture, leadership capacity, and intergenerational collaboration.

### Contributions to the Literature

This study adds to the literature on rural health workforce development in several ways. First, it offers a Philippine-based account of young rural healthcare workers’ experiences, addressing a field still largely informed by evidence from high-income settings. Second, it foregrounds community-integrated mentorship as a culturally embedded learning process, extending current work on rural mentorship beyond formal programmatic approaches. Third, it brings generational adaptation into focus, providing concrete insights on how rural organizations can align with the values and expectations of Generation Z health workers. Finally, it proposes a holistic conceptual model that links structural, relational, and cultural dimensions of organizational support to the sustainability of the rural health workforce in low- and middle-income contexts.

### Strengths and Limitations

A key strength of this study is its focus on early-career rural healthcare workers, whose perspectives are critical for long-term workforce planning but often underrepresented in empirical research. The qualitative design enabled rich, nuanced accounts of organizational support, while sampling across multiple professions and municipalities enhances the potential transferability of the findings.

However, the modest sample size, though appropriate for qualitative inquiry, limits statistical generalizability, and the concentration on three municipalities in a single region means experiences elsewhere in the Philippines may differ. Social desirability bias may also have shaped participants’ descriptions of leadership and organizational culture, despite efforts to ensure confidentiality. Future studies could use longitudinal or mixed-methods designs, larger or more diverse samples, and cross-regional comparisons to deepen and test these insights over time.

## CONCLUSION

This study demonstrates that young rural healthcare workers thrive when organizational structures and practices align with their developmental needs, cultural context, and generational expectations. Community integrated mentorship, supportive work environments, professional growth opportunities, cultural competence, and generational adaptability collectively form a robust foundation for rural workforce sustainability. Strengthening these elements can help rural health systems attract, develop, and retain the next generation of healthcare professionals—ultimately improving health outcomes in underserved communities.

## DECLARATIONS

### Ethics Approval and Consent to Participate

This study was conducted in accordance with institutional and national ethical guidelines for research involving human participants. Ethical approval was obtained from the Philippine Region 1 Ethical Review Committee (Protocol No. NURDD826A 2026). All participants were informed of the study objectives, procedures, potential risks, and their rights prior to participation. Written informed consent was obtained from all participants. Confidentiality, anonymity, and voluntary participation were strictly upheld throughout the research process.

### Consent for Publication

Not applicable. The manuscript does not contain any identifiable personal data.

### Availability of Data and Materials

The qualitative datasets generated and analyzed during this study are not publicly available due to confidentiality agreements with participants. De identified excerpts may be made available from the corresponding author upon reasonable request.

## Competing Interests

The authors declare that they have no competing interests.

## Funding

This research did not receive any specific grant from funding agencies in the public, commercial, or not for profit sectors.

## Authors’ Contributions

Dr. Fernan Torreno conceptualized the study, developed the methodology, conducted data collection, performed the formal analysis, and drafted the initial manuscript. Frincess Flores contributed to validation, supervision, and critical revision of the manuscript for important intellectual content. Both authors reviewed and approved the final version of the manuscript.

## Data Availability

All data underlying this study, including de‑identified interview transcripts and analytic codebooks, are not publicly available due to confidentiality concerns but can be obtained from the corresponding author upon reasonable request and subject to ethical approval.

## Acknowledgements

The authors extend their sincere appreciation to the healthcare workers who generously shared their time and experiences. The authors also acknowledge the support of the local government units and rural health units that facilitated data collection.

## Notes

### Competing Interest Statement

The authors have declared no competing interest.

### Funding Statement

NO FUNDING

### Author Declarations

Ethical approval was obtained from the Philippine Region 1 Ethical Review Committee (Protocol No. NURDD826A-2026).

